# Automated identification of unstandardized medication data: A scalable and flexible data standardization pipeline using RxNorm on GEMINI multicenter hospital data

**DOI:** 10.1101/2022.02.16.22268694

**Authors:** Riley Waters, Sarah Malecki, Sharan Lail, Denise Mak, Sudipta Saha, Hae Young Jung, Fahad Razak, Amol Verma

## Abstract

**Objective:** Patient data repositories often assemble medication data from multiple sources, necessitating standardization prior to analysis. We implemented and evaluated a medication standardization procedure for use with a wide range of pharmacy data inputs across all drug categories, which supports research queries at multiple levels of granularity.

**Methods:** The GEMINI-RxNorm system automates the use of multiple RxNorm tools in tandem with other datasets to identify drug concepts from pharmacy orders. GEMINI-RxNorm was used to process 2,090,155 pharmacy orders from 245,258 hospitalizations between 2010 and 2017 at 7 hospitals in Ontario, Canada. The GEMINI-RxNorm system matches drug-identifying information from pharmacy data (including free-text fields) to RxNorm concept identifiers. A user interface allows researchers to search for drug terms and returns the relevant original pharmacy data through the matched RxNorm concepts. Users can then manually validate the predicted matches and discard false positives. We designed the system to maximize recall (sensitivity) and enable excellent precision (positive predictive value) with minimal manual validation. We compared the performance of this system to manual coding (by a physician and pharmacist) of 13 medication classes.

**Results:** Manual coding was performed for 1,948,817 pharmacy orders and GEMINI-RxNorm successfully returned 1,941,389 (99.6%) orders. Recall was greater than 98.5% in all 13 drug classes, and the F-Measure and precision remained above 90.0% in all drug classes, facilitating efficient manual review to achieve 100.0% precision. GEMINI-RxNorm saved time substantially compared to manual standardization, reducing the time taken to review a pharmacy order row from an estimated 30 seconds to 5 seconds and reducing the number of rows needed to be reviewed by up to 99.99%.

**Discussion and Conclusion:** GEMINI-RxNorm presents a novel combination of RxNorm tools and other datasets to enable accurate, efficient, flexible, and scalable standardization of pharmacy data. By facilitating efficient minimal manual validation, the GEMINI-RxNorm system can allow researchers to achieve near-perfect accuracy in medication data standardization.

## BACKGROUND AND SIGNIFICANCE

Patient data repositories are growing in size and complexity and have increasing importance in a wide range of research applications ^[1-2]^. Medication data are an essential component of these data repositories and require extensive and accurate standardization to enable reliable and reproducible research. This presents a challenge for large medication databases aggregated from multiple sources that may use varying medication vocabularies, data formats, storage systems, or data quality protocols.

Existing standardization methods for medication data are often inflexible, providing limited support for situations where standardized medication identifiers have low coverage or inconsistent formats ^[3-6]^. They typically do not account for free-text drug fields that may contain abbreviations, spelling variations, or additional information that obscures simple parsing ^[3-6]^. Point-in-time manual mapping operations may be time-consuming or result in human error.

Mapping operations are typically performed with the aid of medication ontologies that record normalized drug descriptions along with their interactions and properties ^[7]^. RxNorm, maintained by the National Library of Medicine ^[8]^, is one such platform that is often used to connect and convert data sources between drug vocabularies ^[9]^. Its publicly available Application Programming Interface (API) ^[10]^ includes tools to: link drug concepts with their related concepts, search drug classifications, convert drug identifiers, and approximately match strings to concepts using a custom Natural Language Processing (NLP) method ^[11]^. These tools have been utilized and evaluated individually for specific goals and drug categories ^[4, 12]^ but there remains unexplored potential in creating a holistic procedure, using multiple tools in tandem, that works on all drug categories and supports research query inputs at any level of granularity using any drug vocabulary input.

Our objective was to define, implement, and evaluate an automated pipeline that makes use of existing RxNorm functionality and other available datasets to standardize medication data collected from multiple healthcare organizations. The system was evaluated for use on the GEMINI database – a large, real-world clinical dataset that aggregates data from hospitals in Ontario, Canada ^[13]^. This study describes a highly flexible, scalable, and accurate approach to drug data standardization that can be replicated in most drug databases internationally with minimal modifications, significantly reducing the need for manual mapping.

## METHODS

### Overall Approach

The GEMINI-RxNorm system consists of two independent modules that can be used together to standardize medication data. The Matching Module extracts all potential RxNorm Concept Unique Identifier (CUI) matches for each unique input containing drug-identifying information in the data. Each CUI identifies a specific RxNorm “concept” - a term denoting a commonly accepted definition of a drug given to RxNorm by a source vocabulary ^[8]^. These concepts have a wide range of granularity. For example, “insulin, isophane” (ingredient), “Humulin” (brand name), and “3 ML insulin isophane, human 70 UNT/ML / insulin, regular, human 30 UNT/ML Pen Injector” (Semantic Clinical Drug/Generic Pack) are each unique concepts with their own CUI, which are considered related by RxNorm. The Matching Module runs with each new ingestion of unprocessed data and stores matches in a cache for efficient querying.

The Query Module provides a user interface for researcher queries, determines which RxNorm concepts to search for, and performs back-matching to return the original pharmacy data that relates to those concepts. The Query Module also enables a final review by a subject matter expert prior to use of data. Both modules were implemented in R and utilized the RxMix API ^[10]^.

We validated the performance of the GEMINI-RxNorm system by comparing its outputs to comprehensive manual mappings assembled by a physician and pharmacist.

### Setting

The GEMINI database collects administrative and clinical data for hospital admissions at multiple sites in Ontario^[13, 14]^. GEMINI data undergo a series of data quality checks as previously described ^[14]^. This evaluation used inpatient pharmacy data for patients admitted to or discharged from the general medicine inpatient service of seven hospital sites ^[13]^. It covers physician medication orders from 245,258 unique admissions between April 2010 and October 2017.

The GEMINI pharmacy data include generic names, brand names, Drug Identification Numbers (DIN, unique identifiers assigned by Health Canada to all drug products) ^[15]^, National Drug Codes (NDC, unique identifiers assigned to drugs by the U.S. Food and Drug Administration) ^[16]^, internal hospital identification codes, route, dose, frequency, and other administrative and prescription information. However, raw data coverage and quality varies greatly because hospitals differ in how they store, extract, and manipulate data prior to GEMINI receiving it. Some hospitals may only record generic names or allow free-text entries with additional prescription instructions. Others may not record NDC or may change their data formats and internal codes over time. The GEMINI-RxNorm system is designed for maximum flexibility, utilizing any available drug-identifying information (key identifier fields) regardless of data format or field coverage.

### Matching Module

#### Step 1. Preprocessing

The matching module (Figure 1) began by extracting all raw data from the key identifier fields (Table 1). Each field was then preprocessed in a different way. NDC data was searched for 10 or 11 digit sequences, ignoring separations by symbols or additional lettering. Sequences below 10 or above 11 digits were nullified to avoid non-NDC identifiers that were mistakenly entered in this field. Similarly, DIN data was searched for 8 digit sequences after removing symbols and letters. Any sequence below 8 digits was padded with leading zeros as DINs are often entered without them in the GEMINI data.

**Figure 1.**
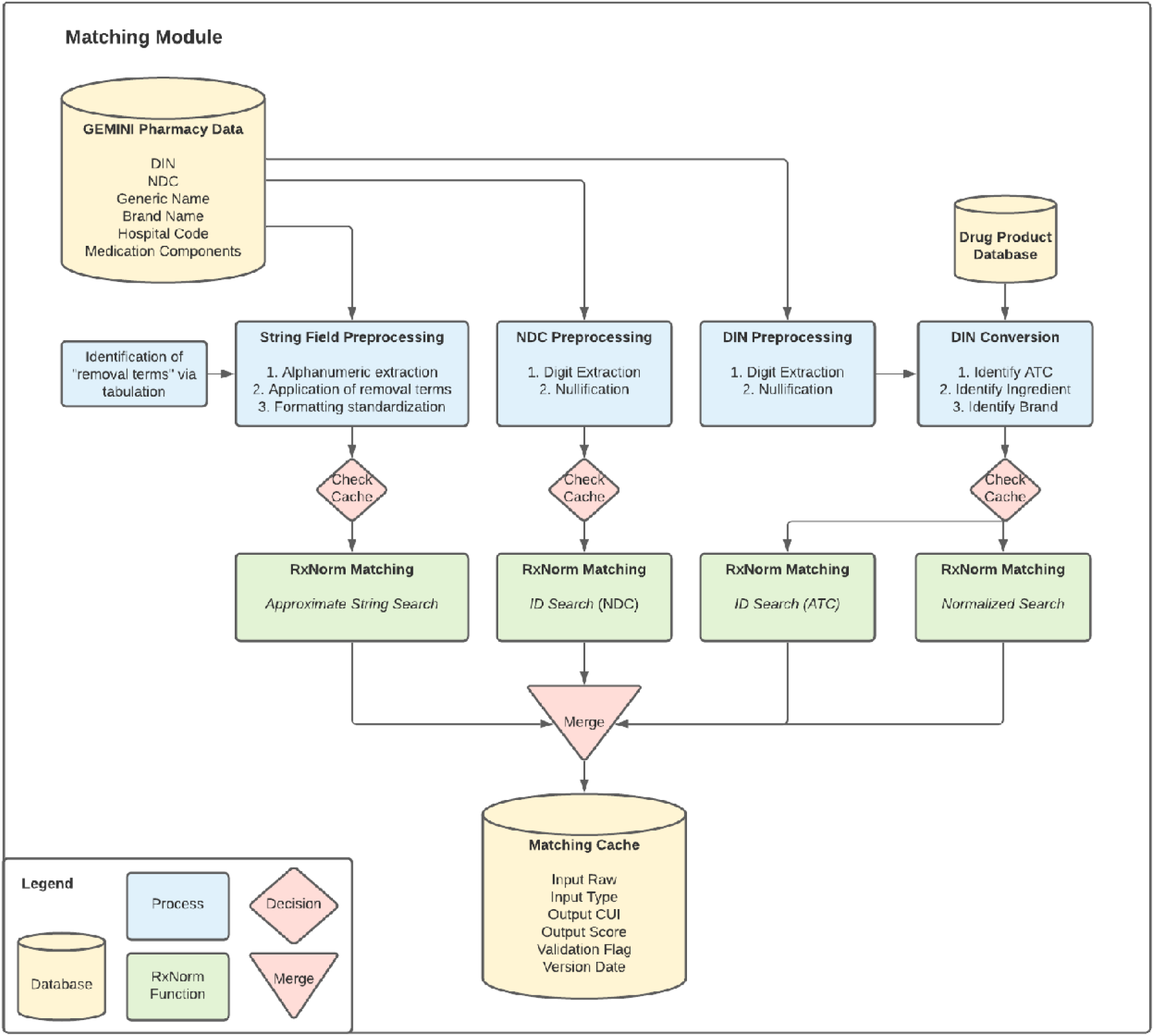
Matching Module The Matching Module matches pharmacy order data to potential RxNorm drug concepts. Pharmacy order data are preprocessed according to their data types (String, NDC, or DIN). A store of outputs called the Matching Cache is checked to see if the data has been encountered before, and if not, an appropriate RxNorm tool is used to determine the matching drug concept identifiers (CUI). All outputs are stored in the Matching Cache along with additional information such as the score indicating how well the data matches each CUI.

**Table 1.** Key Identifier Fields used in the GEMINI database. This table lists the fields that often contain drug identifying information (Key Identifier Fields) in the GEMINI database. Other medication repositories may contain different Key Identifier Fields, but GEMINI-RxNorm will support any number as long as they can be categorized into text or identifier data types.

| **Key Identifier Field** | **Data Type** |
| --- | --- |
| Generic Name | Text |
| Brand Name | Text |
| Drug Identification Number (DIN) | Identifier |
| National Drug Code (NDC) | Identifier |
| Internal Hospital Code | Text |
| Medication Components | Text |

The remaining string fields were preprocessed to only retain strings relevant to RxNorm concepts. Alphanumeric sequences were extracted and symbols discarded excluding those commonly found in RxNorm concept names (“.”, “%”, “-”, “/”). A list of “removal terms” was then identified by tabulating all individual words in the string fields, and manually flagging high-frequency terms that were not relevant to drug identification (e.g. “(NF)” indicating non-formulary, and “(NP)” indicating “Not Preferred”). This is an optional preprocessing step to improve match scores, but it only needs to be done once and can be applied to all string data. Additional custom formatting steps were undertaken such as collapsing whitespace. To retain flexibility for future string formats, no other preprocessing was done. RxNorm Approximate String Search is designed to handle specific wording often found in pharmacy data including dosage and units ^[11]^.

#### Step 2. Concept Matching

Processed data from fields that potentially contained drug-identifying information such as names or IDs, referred to as “key identifier fields” (Table 1), were matched to potential RxNorm CUIs using different tools provided by the RxNorm API. NDC data was inputted directly into the RxNorm *ID Search* function. All string fields were sent through the *Approximate String Search* function. This function supports all RxNorm concept types including generic and brand names.

As RxNorm only supports US drug vocabularies, we converted the processed DINs into identifiers that are supported: Anatomical Therapeutic Chemical (ATC) code, ingredient name, and brand name. This was accomplished by assembling the required fields from the most current Drug Product Database provided by Health Canada ^[17]^ then searching for the identifiers related to the processed DINs. ATC matches were inputted into RxNorm’s *ID Search* while ingredient and brand name matches were inputted into the *Normalized String Search* function. It is important to note that ATC searches lose route information when converted to CUI.

#### Step 3. Building the Matching Cache

A cache of each raw input and its CUI outputs were saved as a database with fields to store the Rx-Norm-defined match score, manual validation flag, and the date the cache was built. This ensured replicability in matching, enabled manual adjustment of matching results, and eliminated the need for re-processing the entire raw data each time a query was made. The output of the *Approximate String Search* can yield multiple potential CUIs along with a match score out of 100 that indicates how closely the input matches the concept. Matches made using the DIN or NDC inputs were assigned a match score of 100 because these were explicit conversions. All of these potential outputs and their match scores were saved.

### Query Module

#### Step 1. Finding Concepts related to a Query

The query module (Figure 2) is an interface for researchers to identify pharmacy data of interest by allowing them to search using any RxNorm supported classification (e.g. Anatomical Therapeutic Chemical (ATC) classifications). For example, a researcher may want to extract all pharmacy records related to diabetes drugs. The user enters keyword(s) of interest (e.g. “diabetes”), and the Query Module returns a list of valid concept names using RxNorm’s *Normalized String Search, Spelling Suggestions, ID Search, All Classes*, and *Class Members* functions^[10]^. After the user confirms all valid concepts of interest, the module uses RxNorm’s *Get Related by Type* function to discover all other directly related concepts. This step ensures that all brand, generic names, and dosage variations of the selected drugs are also searched. For example, a search for “metformin” would also return rows where the only identifying information is the metformin brand name “Glucophage”.

**Figure 2.**
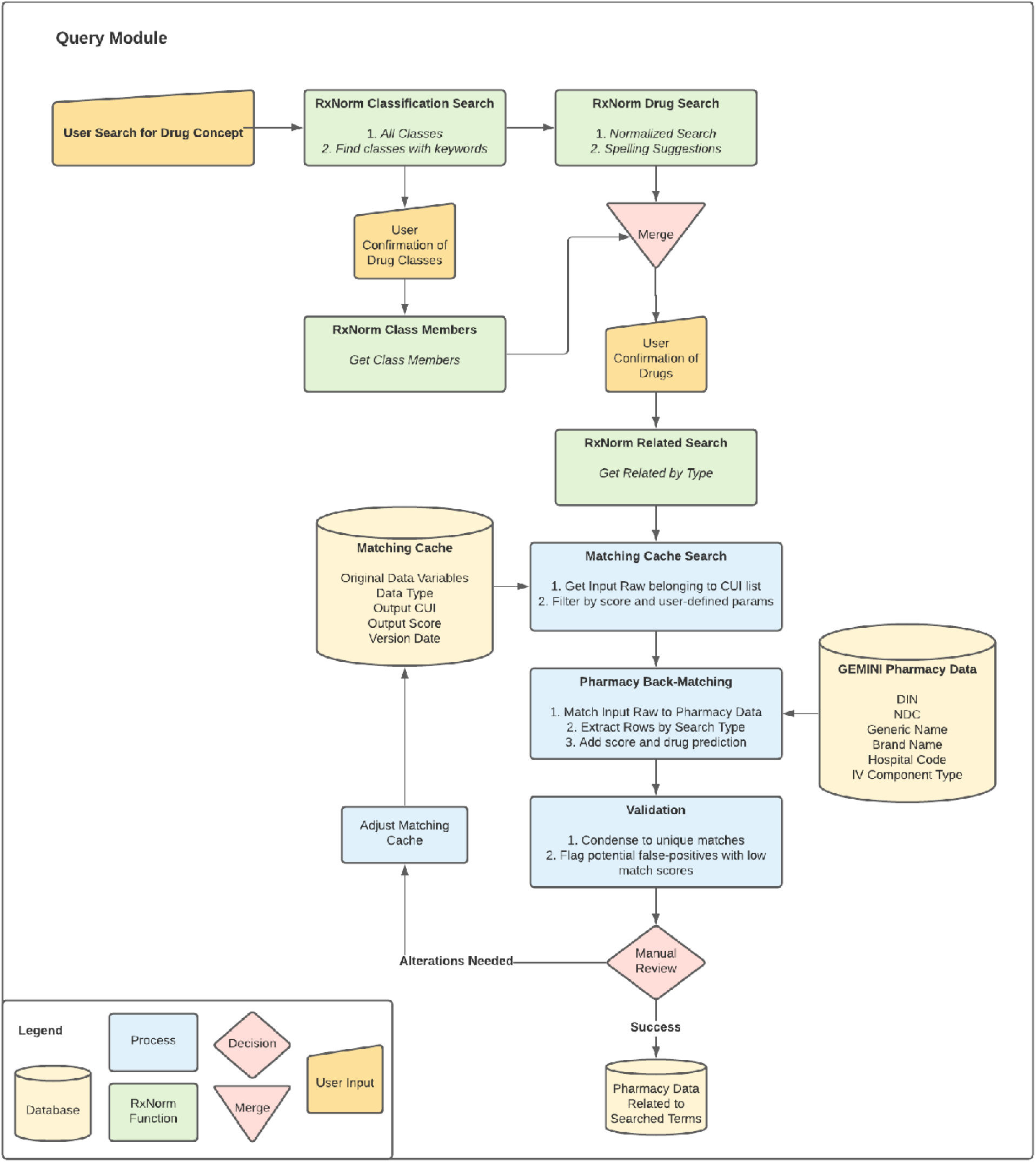
Query Module The Query Module is a researcher interface for retrieving pharmacy orders after the Matching Cache has been created by the Matching Module. The user inputs a list of drug keywords related to the pharmacy orders they want returned (eg. generic names, brand names, ATC codes). RxNorm is then used to determine the list of concept identifiers (CUI) that relate to the user input. The pharmacy orders with potential matches to the CUI list are then identified through a process of back-matching. Outputs are condensed and validated by manual review. False positives may be altered in the Matching Cache to improve precision in future queries.

#### Step 2. Matching Concepts to the Pharmacy Data

With the final concept list constructed, the corresponding CUIs are searched in the matching cache to return the GEMINI pharmacy data variables that match them. These data variables are then back-matched in the GEMINI pharmacy data repository, depending on their field type, to return the rows of original GEMINI pharmacy data that match the search (i.e. orders that included diabetes drugs). The results cache may be further limited to only return matches above a specified match score, between certain dates, or belonging to certain patient encounters or hospital sites.

#### Step 3. Output Flagging

Data users may want to manually check standardized medication data for errors before conducting analyses. It is much less labour intensive to check for “false positive” results (ie. incorrect matches) than “false negatives” (ie. missed matches), because the latter requires manually searching the entire dataset whereas the former only requires checking the suggested matches for correctness. Thus, the GEMINI-RxNorm system was designed to maximize recall (sensitivity), allowing researchers to focus only on manually flagging false positive results. Other applications could easily adjust the system to optimize for other balances of precision and recall if a human in the loop is not desired.

To allow a person to easily identify false positive results, outputted rows are condensed into unique combinations of key identifier fields and their predicted CUI matches. In our case, a pharmacist manually reviewed the matches and removed false matches. Any pharmacist-flagged false matches were removed from the matching cache so that they would not be made on future runs of the system.

### Validation of the GEMINI-RxNorm System

To establish a gold standard of drug mapping, key identifier fields covering 1,948,817 total pharmacy orders for a subset of commonly-used medication classes (Table 2) were manually mapped and validated by a physician (3rd year internal medicine resident) and pharmacist. For each drug category, the physician performed line-by-line manual coding using a master file of pharmacy data for each hospital site. Variables used to code drugs into categories included brand name, generic name, DIN and route information where available. Unique combinations of variables were displayed as rows and lines for mapping. The number of occurrences of each unique combination was also provided to offer additional context with respect to the most common medication orders.

**Table 2.** Drug Classifications used for Validation This table defines the input drugs used in each query. Combination products were listed separately. Items that list an Anatomical Therapeutic Chemical (ATC) code indicate that the entire list of generic drugs as specified by ATC were used as input.

| **Drug Classification** | **Drugs included** |
| --- | --- |
| Insulin | Anatomical Therapeutic Chemical (ATC): A10A |
| Non-insulin glucose lowering drugs | ATC: A10B |
| Vitamin K Antagonists | ATC: B01AA |
| P2Y12 Inhibitors | clopidogrel, ticagrelor, prasugrel |
| Ace inhibitors/Angiotensin II Receptor Blockers | ATC: C09A, C09C |
| Benzodiazepines | ATC: N05BA |
| Direct-acting Oral Anticoagulants | ATC: B01AF |
| Antipsychotics | ATC: N05A |
| Dementia Medications | donepezil, memantine, galantamine, rivastigmine |
| Furosemide | furosemide (Lasix) |
| Steroids | dexamethasone, hydrocortisone, methylprednisolone, prednisolone, prednisone, deflazacort, cortisone |
| Puffers (limited to inhalation route) | albuterol, salbutamol, levalbuterol, terbutaline, Proventil, ProAir, AccuNeb, pirbuterol, ventolin, formoterol, salmeterol, indacaterol, arformoterol, olodaterol, orciprenaline, ipratropium, umeclidinium, glycopyrrolate, glycopyrronium, tiotropium, aclidinium, budesonide, fluticasone, beclomethasone, ciclesonide, mometasone, albuterol/ipratropium, albuterol/ipratropium, umeclidinium/vilanterol, olodaterol/tiotropium, glycopyrronium/indacaterol, formoterol/glycopyrronium, aclidinium/formoterol, tiotropium/olodaterol, fluticasone/salmeterol, fluticasone/vilanterol, budesonide/formoterol, beclomethasone, |
|  | salmeterol, formoterol/mometasone, beclomethasone/formoterol/glycopyrronium, fluticasone/umeclidinium/vilanterol, budesonide/glycopyrronium/formoterol, ibudilast, montelukast, pranlukast, zafirlukast, roflumilast, aminophylline, doxofylline, theophylline |

The GEMINI-RxNorm system was run once on the same subset of manually-coded data. It queried the data for the same drug names as the manual mappings and had no restrictions on the minimum match score required to make a match. No manual revisions were made to the GEMINI-RxNorm outputs to ensure that only the automated standardization procedure would be evaluated. We then calculated the F-measure, precision, and recall of the outputs by comparing to the gold standard. Analyses were performed using R version 4.0.1. Research ethics board approval for this study was obtained from all participating sites.

## RESULTS

The GEMINI-RxNorm system performed matching on 2,090,155 medication orders, occurring during 245,258 hospital admissions at 7 hospitals between April 1, 2010 and October 31, 2017. The system categorized the medication orders into 29,249 distinct RxNorm CUIs.

### Validation

The pharmacy-order level results (Table 3) reflect the real-world performance of the system when retrieving individual GEMINI pharmacy data for the given queries. In total, GEMINI-RxNorm successfully returned 1,941,389 of the 1,948,817 (99.62%) manually identified orders. The recall (sensitivity) of the GEMINI-RxNorm system was above 98.5% for all medication classes and the F-measure was above 95% in all drug categories except steroids (91.82%) and antibiotics (90.15%). With minimal manual review to discard false positives, precision of 100.0% can be achieved. The majority of false positives were caused by orders that included drugs with a similar string-name to a drug concept related to a queried drug. For example, “Cipralex” (escitalopram) orders were assigned a 50% match score to the brand name “Ciprodex” leading these orders to be incorrectly matched to the antibiotic ciprofloxacin. Cases such as these could be avoided by setting a minimum match score above 50% but doing so could potentially lower system’s recall (Figure 3). There was a marked improvement in precision at match scores of 50% with relatively little tradeoff in recall. Medication orders that the system could not match were orders where the only drug-identifying information was a Canada-specific drug name such as “Gravol” which RxNorm cannot recognize.

**Table 3.** Validation Results Results of the validation for the 13 drug classification queries. For each query, the pharmacy orders returned by GEMINI-RxNorm (with no limitations on match scores) were compared to the gold standard manual mappings. TP: True Positive; FP: False Positive; FN: False Negative

| **Drug Class** | **Number of Orders** | **F-Measure** | **Precision** | **Recall** |
| --- | --- | --- | --- | --- |
| Insulin | TP: 141349<br>FP: 1910<br>FN: 1580 | 98.8 | 98.7 | 98.9 |
| Non-insulin glucose lowering drugs | TP: 106463<br>FP: 9480<br>FN: 39 | 95.7 | 91.8 | >99.9 |
| Vitamin K Antagonists | TP: 100023<br>FP: 4632<br>FN: 0 | 97.7 | 95.6 | 100 |
| P2Y12 inhibitors | TP: 42720<br>FP: 0<br>FN: 0 | 100 | 100 | 100 |
| Ace inhibitors/Angiotensin II Receptor Blockers | TP: 133398<br>FP: 6548<br>FN: 86 | 97.6 | 95.3 | 99.9 |
| Benzodiazepines | TP: 167577<br>FP: 2268<br>FN: 1465 | 98.9 | 98.7 | 99.1 |
| Direct-acting Oral Anticoagulants | TP: 22970<br>FP: 82<br>FN: 0 | 99.8 | 99.6 | 100 |
| Antipsychotics | TP: 149213<br>FP: 128<br>FN: 287 | 99.9 | 99.9 | 99.8 |
| Dementia Meds | TP: 16262<br>FP: 0<br>FN: 0 | 100 | 100 | 100 |
| Furosemide | TP: 220422<br>FP: 14 | >99.9 | >99.9 | >99.9 |
|  | FN: 52 |  |  |  |
| Steroids | TP: 147470<br>FP: 26286<br>FN: 0 | 91.8 | 84.9 | 100 |
| Puffers | TP: 242622<br>FP: 22232<br>FN: 3261 | 95.0 | 91.6 | 98.7 |
| Antibiotics | TP: 450900<br>FP: 97906<br>FN: 658 | 90.2 | 82.2 | 99.9 |

**Figure 3.**
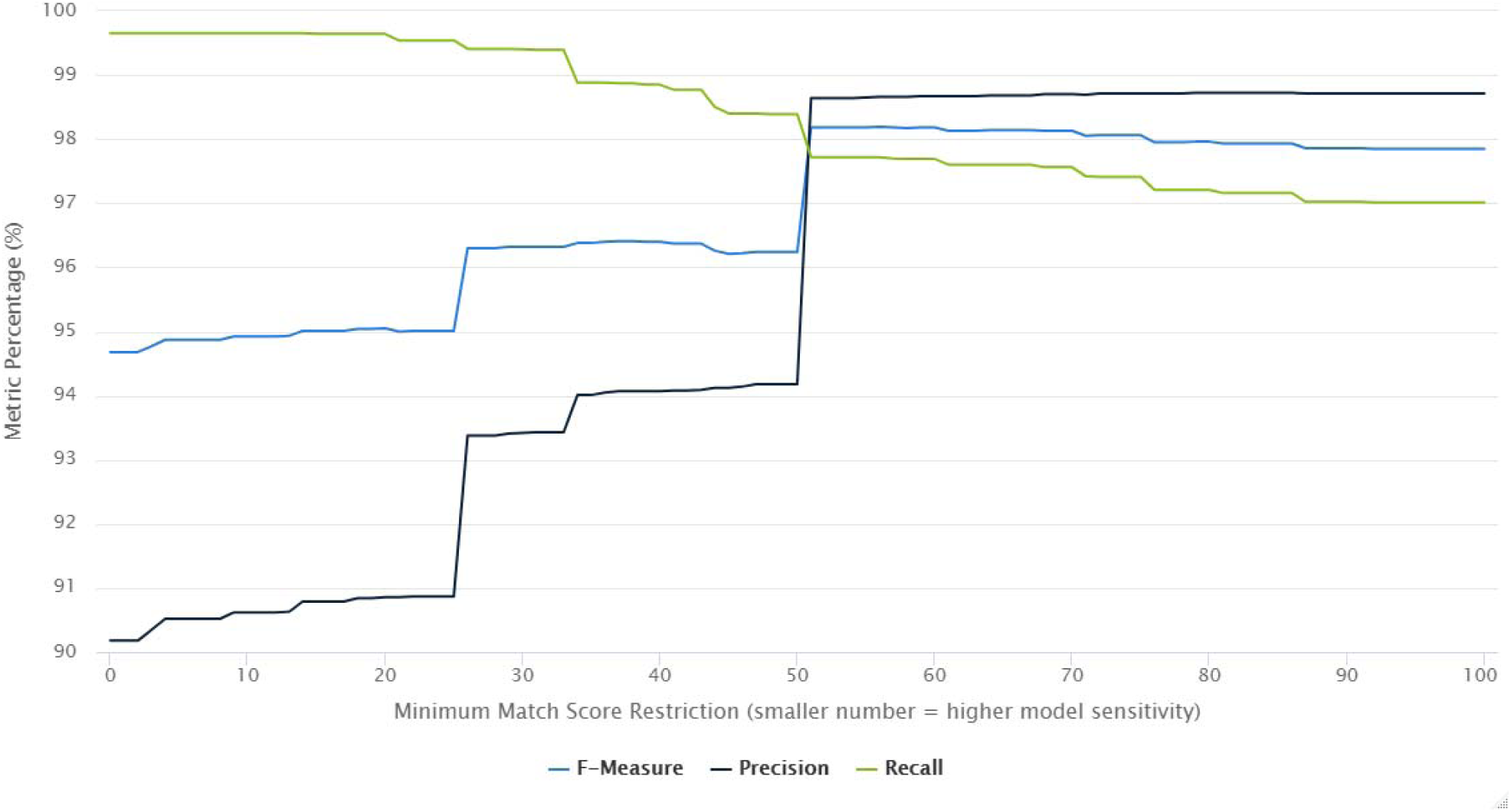
Gemini-RxNorm Performance at Different Match Score Levels This chart displays the performance metrics of GEMINI-RxNorm when run using different “minimum match scores”. The match score is a number between 0 and 100 that RxNorm uses to indicate how closely a free-text string matches an RxNorm concept. The x-axis in this chart represents the minimum match score needed for GEMINI-RxNorm to return the match. The y-axis represents the percentage metrics of F-measure, precision, and recall obtained by comparing the pharmacy orders returned by GEMINI-RxNorm to the 1,948,817 gold standard manually mapped pharmacy orders.

### Time Savings

To illustrate the time saved by the GEMINI-RxNorm system compared to manual medication mapping, we estimated that the time required to manually map a single row of medication data to a drug category was 30 seconds whereas the time required to manually verify suggested medication category matches through GEMINI-RxNorm was 5 seconds per row. To identify insulin medications, a manual reviewer might need to check up to all 2,090,155 pharmacy orders whereas after application of GEMINI-RxNorm, manual review was only required for 662 consolidated rows of data.

## DISCUSSION

This paper describes the development, implementation, and extensive validation of GEMINI-RxNorm, a medication data standardization system that uses a novel combination of RxNorm tools and external datasets. GEMINI-RxNorm demonstrates a flexible approach to medication standardization that can support a variety of input types, data formats, quality, and coverage that may be found when aggregating raw medication orders from multiple sources. The system was compared to manual expert mappings of 13 drug classes from 7 Canadian hospital sites over 8 years. It was found to have recall greater than 98.5% and an F-measure above 90.0% in all classes. GEMINI-RxNorm enabled substantial time savings compared to full manual standardization, reducing the time taken to review a pharmacy order row from an estimated 30 seconds to 5 seconds and reducing the number of rows needed to be reviewed by up to 99.99%, from 2,090,155 rows to 662. Our experience suggests that the GEMINI-RxNorm system can be used independently to efficiently extract and standardize pharmacy data with a high degree of accuracy. With minimal additional manual validation, researchers can achieve nearly perfect accuracy of standardized medication data in multisite patient data repositories.

Data standardization in clinical research repositories is crucial as it can have major impacts on research outcomes and policy decisions. Zhou and colleagues describe an automated method to map the Partners Master Drug Dictionary (MDD) to RxNorm concepts using the MTERMS NLP tool ^[18]^. Similarly, Jiang and colleagues standardized medication information in clinical text using MedEx ^[19]^. These studies focus on point-in-time mappings with single-source inputs. Less has been published on defining scalable frameworks for use in growing research repositories. Other drug data standardization efforts have involved mapping specific drug categories to RxNorm concepts. Klevens and colleagues categorized outpatient antibiotic prescription claims ^[3]^ and Dhavle et al. evaluated 49,997 ambulatory e-prescription claims using RxNorm^[4]^. These studies had a limited range of data inputs and were mapped using NDCs provided in their starting data without support for free-text parsing. We have been unable to find any studies that combine and evaluate several RxNorm tools in tandem with the intention of mapping all drug categories and supporting the continuous standardization that is required by ongoing data collection in growing research repositories. We describe the implementation of RxNorm in a health system outside of the United States, and by defining processes to link Canadian DINs to RxNorm concepts, the GEMINI-RxNorm framework can be easily extended for other international applications. Although optimized for use within the GEMINI dataset, the GEMINI-RxNorm system is flexible and can be implemented in medication repositories with different types of input data as it is designed to extract drug-identifying concepts from a wide range of fields.

Some limitations remain in the GEMINI-RxNorm system. The RxNorm functions that we used do not support non-US drug concepts and therefore, it was not possible to match some non-US medication brand names to RxNorm concepts. This represents an opportunity for future work involving custom natural language processing tools or efforts to expand RxNorm to international medications such as with OHDSI’s RxNorm Extension. Additionally, GEMINI-RxNorm allows users to search using classifications such as ATC, but doing so will not allow for specific medication routes to be searched. GEMINI-RxNorm was able to identify drugs even when abbreviated or obscured among additional information, but it can misidentify similar-sounding drugs and does not take into account a complete view of a drug order that a human may find. For example, some furosemide orders included the string “Hold Lasix for Today”, indicating that Lasix was not ordered. However, GEMINI-RxNorm returned this row in the furosemide query as it only saw the word “Lasix”. Our experience indicates that manual data validation is still necessary to resolve these cases, and thus we designed a module to facilitate this process. Starting with automated standardization, maximizing the system’s recall, and then condensing the outputs for manual review can minimize the manual workload while maintaining data quality. Any false positives flagged by the reviewer can then be removed from the matching database so that future queries do not make the same mismatch. The drug classes we validated were chosen to represent a wide range of medications commonly used in research, but many drug classifications were not validated. We believe that the GEMINI-RxNorm tool is likely to perform with the same excellent recall/sensitivity for most common medications, given its consistent performance across the wide range of classes that were validated.

## CONCLUSION

The GEMINI-RxNorm system is a comprehensive, flexible, scalable, and highly accurate automated pipeline for drug standardization in multisite patient data repositories. Extensive manual validation demonstrates consistently excellent recall and very good precision for medications across a wide range of medication classes. Thus, with limited additional manual validation, the GEMINI-RxNorm system can allow researchers to achieve near-perfect accuracy in medication data standardization.

## Data Availability

Data from this manuscript can be accessed upon request to the corresponding author, to the extent that is possible in compliance with local research ethics board requirements and data sharing agreements

